# Systematic review of the association between ABO blood type and COVID-19 incidence and mortality

**DOI:** 10.1101/2021.04.20.21255816

**Authors:** Yuqing Bai, Zhou Yan, Eleanor J Murray

**Affiliations:** Department of Epidemiology, Boston University School of Public Health, Boston, MA

## Abstract

A large proportion of COVID-19 research has been focused on identifying markers of high-risk individuals. However, this research often fails to consider basic epidemiologic concepts to prevent bias in the design, selection, and analysis of observational data. One suspected marker of risk that has been repeatedly assessed is ABO blood type. Given the ease of measuring this biomarker, it is an appealing target for identifying high-risk individuals. However, this same ease of measurement makes associational research on ABO blood type and COVID prone to a range of common epidemiologic errors. We conducted a systematic review of studies assessing correlations between ABO blood type and COVID incidence, hospitalization, and mortality to determine the quality of evidence these studies provide and whether the overall evidence suggests ABO blood type could provide a useful indicator of COVID risk. We conclude that most existing studies are low quality and suffer from major methodological flaws. The few higher-quality studies which do exist find no association between ABO blood type and COVID outcomes. We conclude that there is no evidence to support the use of ABO blood type as a marker for COVID risk or severity.

**Key Points:**

- There is no sufficient evidence to conclude a biological relationship between ABO blood types and COVID-19 infection or severity.
- Biases of existing research could be avoided by careful study design.

## Introduction

The COVID-19 pandemic has placed a tremendous burden on healthcare and public health systems worldwide. In an attempt to improve pandemic response, there has been sustained research interest in investigating potential biological characteristics which could be used as markers for COVID-19 risk, severity, or SARS-CoV-2 infection status. This so-called ‘risk factor epidemiology’ is appealing because prediction or early identification of individuals who will get sick, need to be hospitalized, or die could allow more targeted public health measures and reduce the pressure on overburdened systems. Unfortunately, risk factor epidemiology has long been criticized for confusing correlation and causation and leading to misleading and incorrect decision-making (Huitfeldt, 2016). Importantly, risk factor epidemiology is not designed to produce reliable and reproducible outcome predictions and making those predictions by any method is challenging. In fact, even state-of-the-art prediction modelling approaches that have been applied to COVID have not succeeded in this goal (Wynants et al., 2020).

Despite this, a number of high-profile studies on ABO blood type as a risk factor for COVID-19 outcomes have drawn the attention of the public. In particular, a genome wide association study (GWAS) conducted by Ellinghaus et al. (2020) showed a positive association between Type A blood group and risk of COVID-19 infection, and a negative association between Type O blood group and risk of COVID-19 infection. Several other studies seemed to support these findings, and this confluence of study results has been used as evidence to suggest the existence of a true biological relationship between ABO blood type and COVID-19 risk or severity. This has led to a (incorrect) belief among members of the public that individuals with Type O blood are not at risk of COVID-19 or cannot contract SARS-CoV-2.

Although it may be tempting to conclude agreement between study results suggests strong evidence in support of the use of ABO blood type to predict COVID-19 outcomes, this is not necessarily correct. The number of studies returning a given result is not in itself a reasonable assessment of the validity of that result, because studies which repeat the same methodological errors can return the same erroneous conclusions. Importantly, several additional studies of ABO blood type and COVID risk have found no association between blood type and risk. This suggests that a deeper look is needed to understand whether a true relationship exists or whether the relationship should be attributed to bias.

Investigation of ABO blood type and COVID-19 necessarily requires the use of observational data --that is, blood type cannot be randomly assigned to individuals, so we must observe COVID-19 outcomes in individuals who happen to have particular blood types. Observational research is challenging to do well, because of potential sources of bias like confounding, selection bias, and measurement error. These sources of bias are well-characterized in the epidemiologic literature, and have been discussed at length with respect to COVID-19 (Griffith et al., 2020). However, many COVID-19 studies fall prey to biases which could be avoided by more careful study design or analytic plans.

Here, we review the literature on COVID-19 and ABO blood type to assess the quality of the evidence for or against a relationship between ABO blood type and COVID-19 risk and severity.

## Methods

We searched PubMed using the following search terms: (“ABO blood-group system” [mesh] OR ABO blood type OR ABO blood group OR ABO) AND (“COVID-19” [Supplementary Concept] OR COVID-19 OR SARS-CoV-2 OR coronavirus*) for articles published before November 2020. In total, our search returned 87 articles. Additionally, 2 additional articles not in PubMed were identified from a Google search (Alkout & Alkout, 2020; Arac et al., 2020). Titles and abstracts of all identified studies were reviewed and resulted in 59 potentially relevant articles. For our review we were interested in epidemiologic, population-level studies that investigated the relationship between ABO blood groups and COVID-19 infection status or other COVID-19 outcomes, therefore we excluded studies which focused on other aspects of a possible relationship between COVID-19 and ABO blood type. After reviewing the full text of each article, we excluded 14 review papers, 8 articles discussing possible molecular mechanisms without including epidemiologic data, and 13 articles focusing on clinical applications or blood transfusions. Our final analysis includes 24 epidemiologic studies identified from our review process. The screening process for articles in PubMed search is summarized in Figure 1.

**Figure 1.**
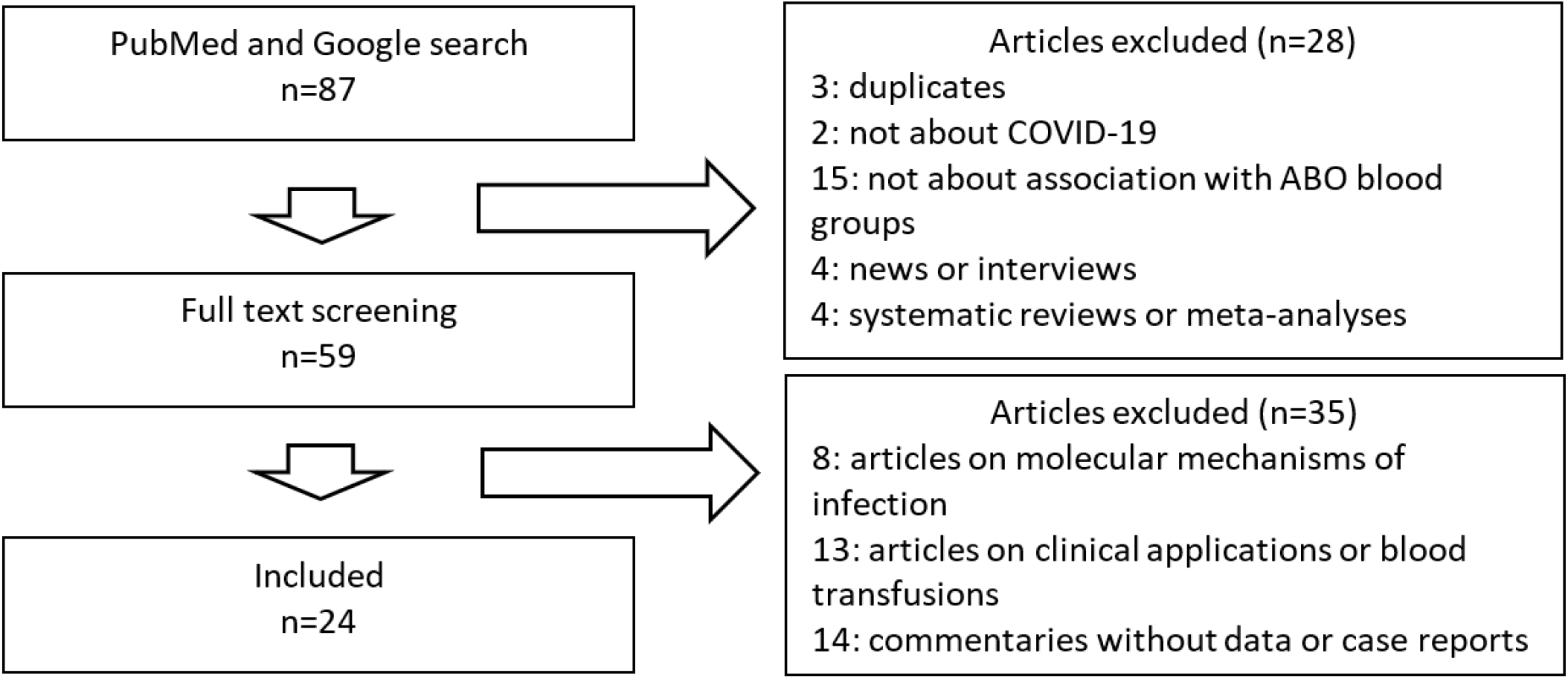
Article screening process.

For each of the 24 final studies, we reviewed the full text to identify potential methodological issues. We used the ROBINS-I (Risk of Bias In Non-randomized Studies - of Interventions) tool by Sterne et al. (2016) to assess the risk of bias level in each article. We chose this tool because discussions of the relationship between ABO blood group and COVID-19 risk suggest the hypothesis under study is about a causal effect. The following domains of bias described in Sterne et al. (2016) were considered for all studies that were reviewed: bias due to confounding; bias in selection of participants into the study; bias in classification of interventions; bias due to missing data; bias in the selection of reported results. One important caveat in the application of this risk of bias tool is that ABO blood group is not an intervention. We therefore did not assess the category “bias due to deviations from intended interventions”. In addition, most studies did not include sufficient detail for us to assess the potential for measurement error in COVID-19 outcomes.

## Results

### Article Screening

The 24 articles we reviewed include 7 retrospective cohort studies, 13 case-control studies, 2 ecological studies, and 3 cross-sectional surveys (Table 1). The articles include 6 studies based in Europe, 6 studies in North America, 5 studies in China (3 were in Wuhan only), 4 studies in Asia excluding China, 1 study in Africa and 2 ecological studies that were conducted on multiple nations. 19 (79.17%) out of the 24 articles assessed risk for infection, 5 (20.83%) studies assessed risk for any COVID symptoms, 2 (8.33%) assessed risk of hospitalization after infection, 8 (33.33%) studied the type of care received such as ICU admission or intubation, and 12 (50.00%) assessed the risk of death due to COVID.

**Table 1:**
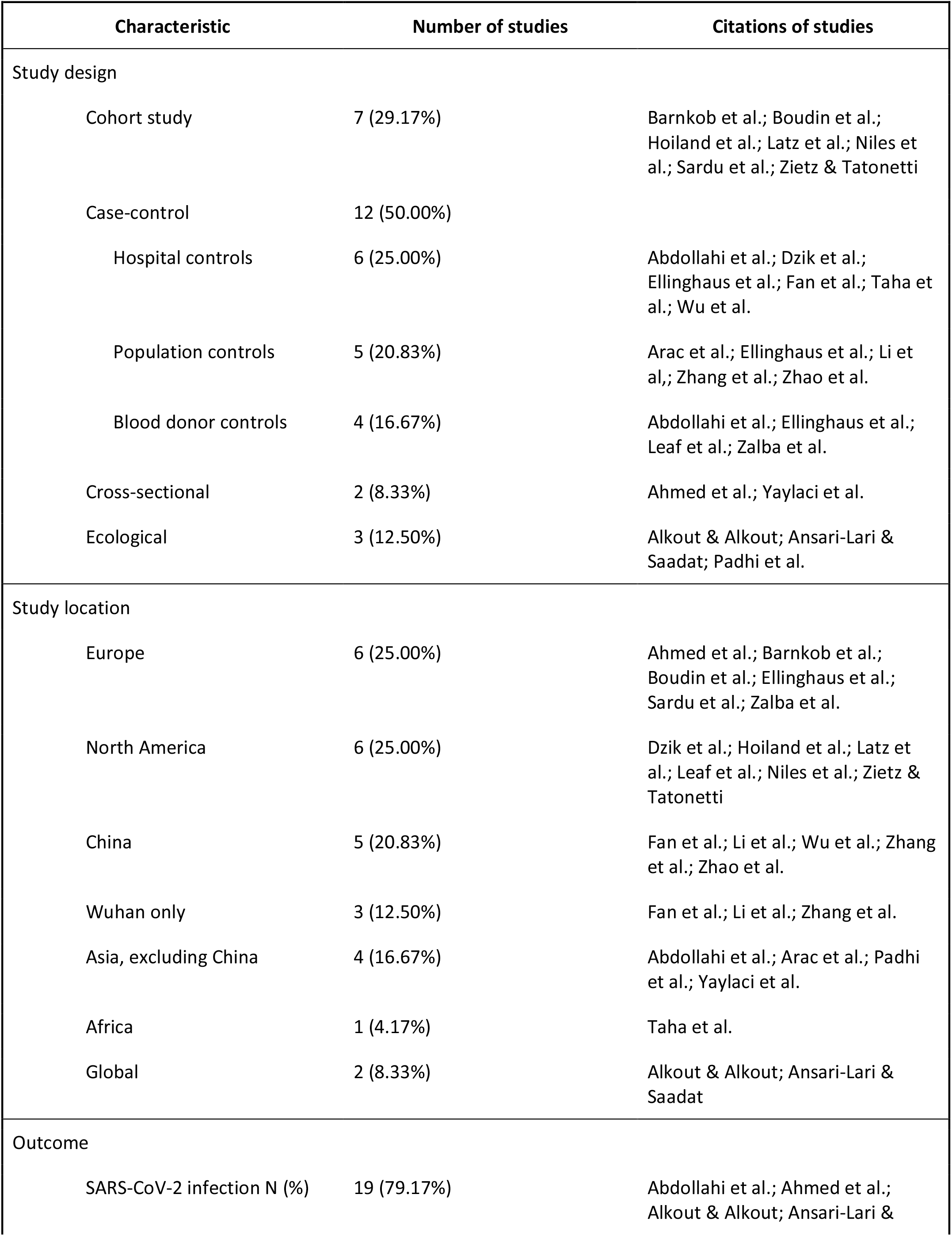
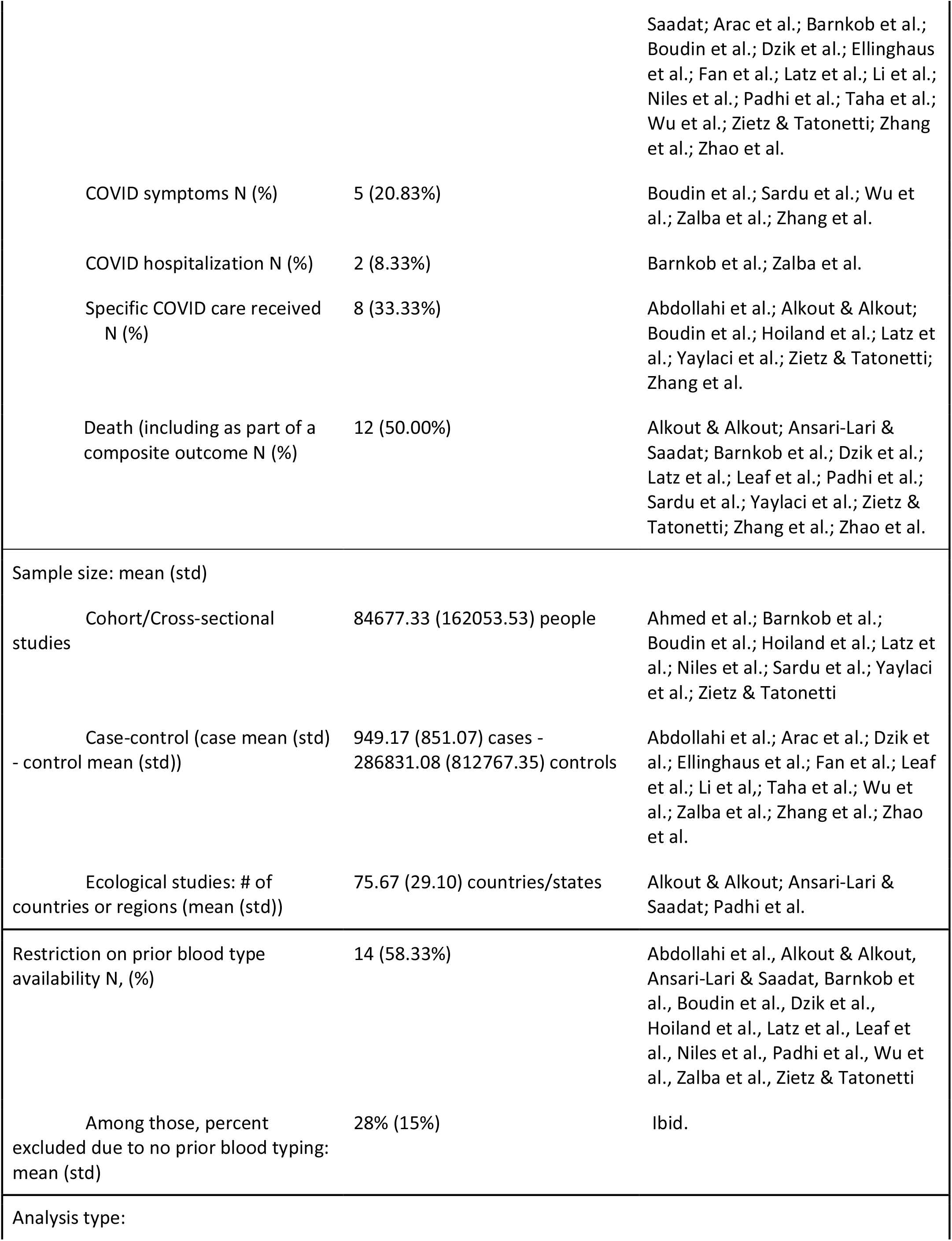
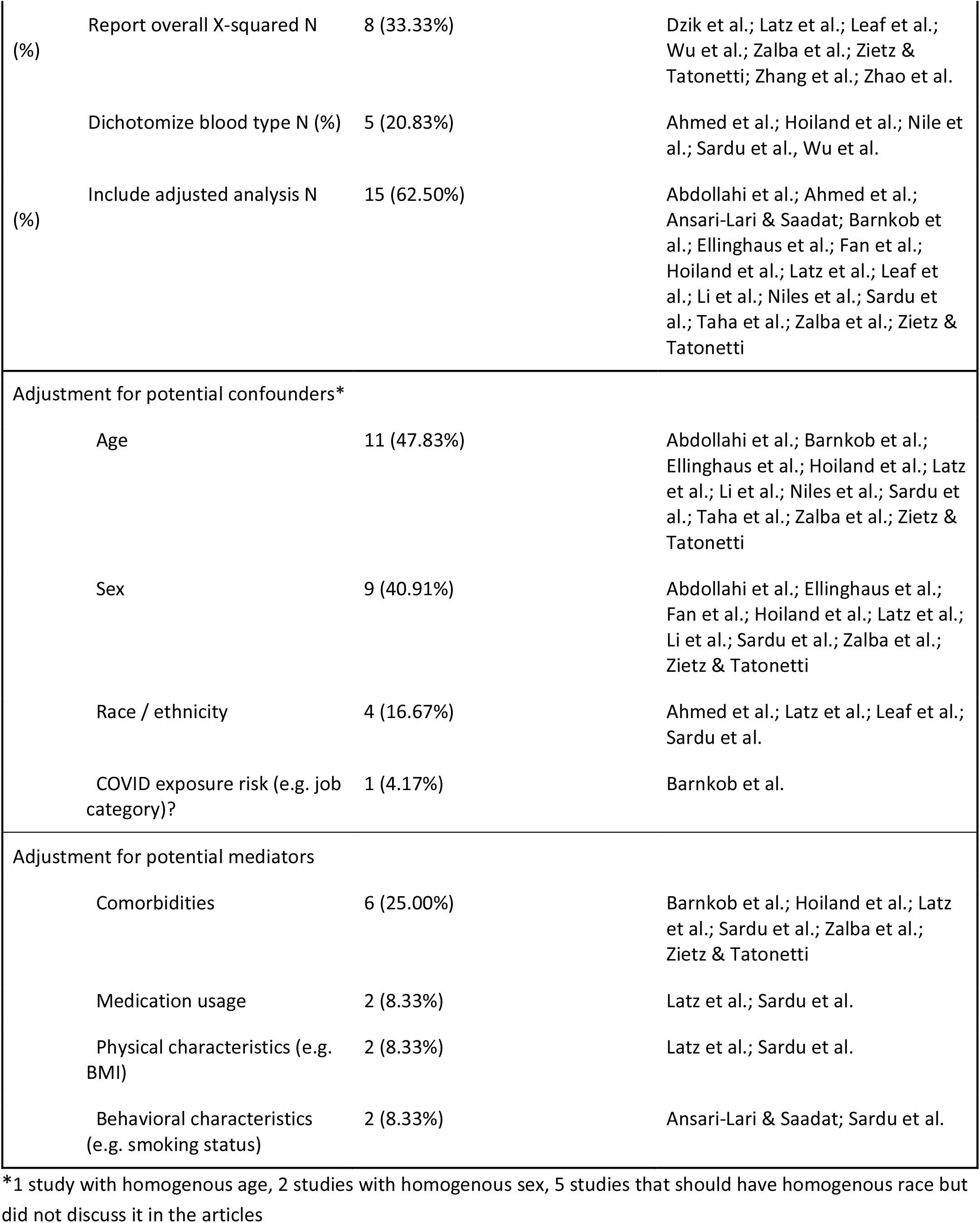
Summary of studies.

Based on the ROBINS-I tool, we concluded that 12 of the 24 articles (50%) had a serious overall risk of bias, and 10 (42%) articles with a moderate overall risk of bias (Table 2). One article (4%) had a low risk of bias (Boudin et al., 2020), and a final article was judged to have low risk of bias for the primary outcome but moderate risk of bias for all other comparisons (Zhang et al., 2020). A summary of issues with each study is included in the Appendix.

**Table 2.** Summary of ROBINS-I review results.

|  | Abdollahi et al. | Ahmed et al. | Alkout & Alkout | Anderson et al.* | Ansari-Lari & Saadat | Arac et al. | Barnkob et al. | Boudin et al. | Dzik et al. | Ellinghaus et al. | Fan et al. | Hoiland et al. | Latz et al. |
| --- | --- | --- | --- | --- | --- | --- | --- | --- | --- | --- | --- | --- | --- |
| **Bias due to confounding** | Moderate | Moderate | Serious | Low | Moderate | Serious | Low | Low | Moderate | Moderate | Moderate | Moderate | Serious |
| **Bias in selection of participants into the study** | Moderate | Low | Moderate | Moderate | Moderate | Moderate | Serious | Low | Moderate | Moderate | Serious | Serious | Serious |
| **Bias in classification of interventions** | Low | Low | Moderate | Low | Moderate | Low | Low | Low | Low | Low | Low | Low | Low |
| **Bias due to missing data** | unclear | unclear | unclear | unclear | unclear | unclear | unclear | Low | unclear | Low | unknown | Moderate | Low |
| **Bias in measurement of outcomes** | Low | unclear | Serious | Low | Serious | Low | Low | Low | Low | Low | Low | Low | Low |
| **Bias in selection of the reported result** | Low | Moderate | Low | Low | Serious | Serious | Low | Low | Low | Low | Low | Low | Low |
| **Overall** | Moderate | Moderate | Serious | Low to Moderate | Serious | Serious | Serious | Low | Moderate | Moderate | Serious | Serious | Serious |

\*Recent article, added following systematic search

**Table 2. Summary of ROBINS-I review results (cont.)**
|  | Leaf et al. | Li et al. | Niles et al. | Padhi et al. | Sardu et al. | Taha et al. | Wu et al. | Yaylaci et al. | Zalba Marcos et al. | Zhang et al. | Zhao et al. | Zietz & Tatonetti |
| --- | --- | --- | --- | --- | --- | --- | --- | --- | --- | --- | --- | --- |
| **Bias due to confounding** | Low | Low | Moderate | Serious | Moderate | unclear | Low | unclear | Low | unclear | unclear | Moderate |
| **Bias in selection of participants into the study** | Serious | Moderate | Moderate | Serious | Low | Serious | Serious | unclear | Low to Moderate | Low to Moderate | unclear | Serious |
| **Bias in classification of interventions** | Low | Low | Low | Low | Low | Serious | Low | unclear | Low | Low | Low | Low |
| **Bias due to missing data** | Low | unclear | Low | unclear | unclear | unclear | Moderate | unclear | unclear | unclear | unclear | unclear |
| **Bias in measurement of outcomes** | Low | Low | Low | Low | Low | Low | Low | Low | Low | Low | unclear | Low |
| **Bias in selection of the reported result** | Low | Moderate | Low | Low | Moderate | Low | Low | Moderate | Low | Moderate | Moderate | Low |
| **Overall** | Serious | Moderate | Moderate | Serious | Moderate | Moderate | Serious | Serious | Moderate | Low to Moderate | Serious | Serious |

As we finalized our manuscript, a large case-control study was published (Anderson et al, 2021). This article was added to our ROBINS-I table and a summary is included in the Appendix. We concluded that the risk of bias in this article was low to moderate.

### Highest-quality studies

Of the 24 studies identified, the two studies with least bias were Boudin et al. which we judged to have a low risk of bias, and Zhang et al. which we judged to be low risk of bias for the primary outcome --a comparison of ABO blood types between COVID survivors and non-survivors. Both of these studies reported no association between ABO blood type and their primary COVID outcomes.

In addition, Anderson et al was judged to be low to moderate bias and reported no association between ABO blood type and any COVID outcome. The impact of confounding is likely minimal due to the fairly homogenous population, but it is unclear how many individuals were excluded due to missing ABO blood type information.

### Reported associations

Overall, 19 of the 24 (79%) studies reported a significant association between at least one ABO blood type and at least one COVID-19 outcome, while 4 (17%) of studies found no association and 1 study found contradictory city-specific results but null pooled results (Zhao et al., 2020). Of the 19 articles that found a significant association, the results were contradictory. Just over half (n = 15; 62.50%)) found an improved outcome (no infection, decreased severity, no death, etc.) among Type O subjects, and about half (n = 13; 54.17%) found worsened outcome (infection, higher severity, death, etc.) among Type A subjects. However, only 8 (33.33%) studies reported both better outcomes for individuals with Type O blood and worse outcomes for individuals with Type A blood at the same time. 4 (16.67%) articles found better outcomes for Type B subjects while 2 (8.33%) found worse outcomes for Type B. Finally, 3 (12.50%) articles found worse outcomes among Type AB subjects while 1 (4.17%) found better outcomes among Type AB.

### A proposed causal diagram

Figure 2 shows a proposed causal directed acyclic graph (DAG) (Hernan & Robins 2020) depicting our best understanding of potential relationship between ABO blood group and COVID-19 outcomes. The DAG also includes possible important confounders, mediators, and sources of bias, as well as factors which determine inclusion or exclusion in the 24 existing studies. We used this DAG to evaluate the potential for bias due to confounding or selection.

**Figure 2.**
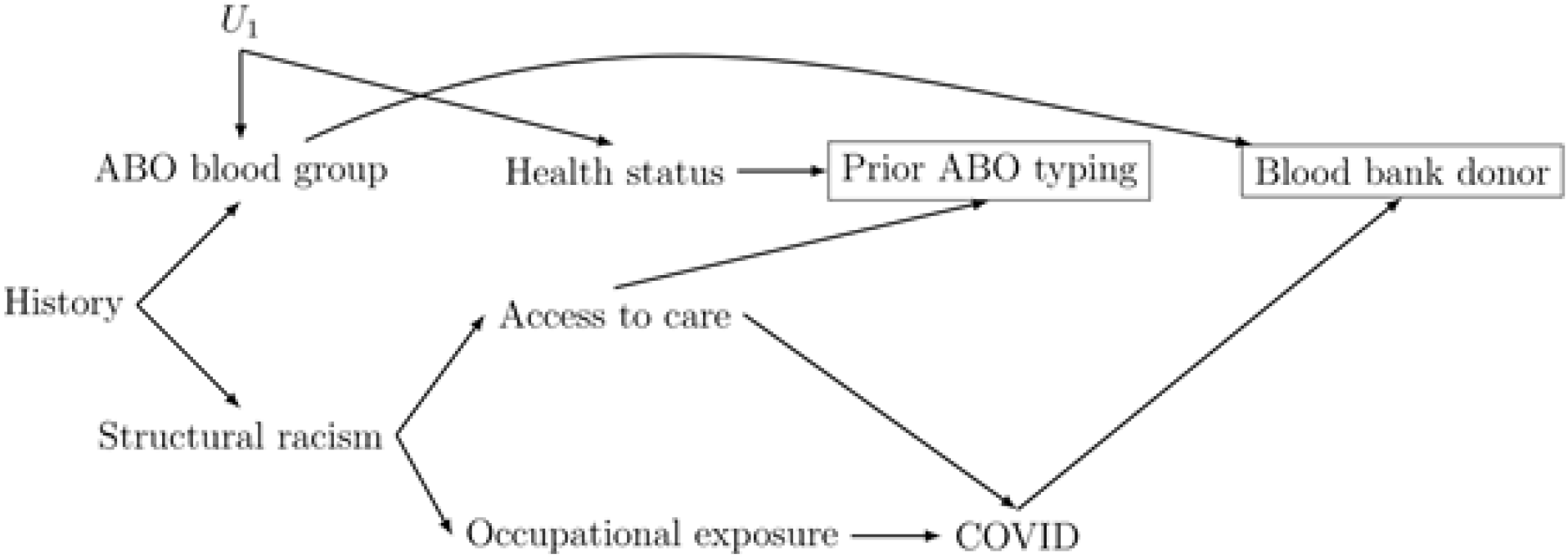
Directed acyclic graph demonstrating potential bias pathways in studies of ABO blood group and COVID-19. Prior ABO typing and blood bank donor status are required inclusion variables in a number of the 24 ABO blood type and COVID-19 outcomes studies, and this restriction is indicated by a box around those nodes. When blood bank donor status is used, it is for identifying control participants and thus a relationship exists between COVID-19 outcomes and blood bank donor status by design despite the fact that this relationship is not biological.

In our DAG, the key confounding variables for ABO blood group and COVID-19 are historic and genetic factors, representing the fact that while an individual’s genome is a random permutation of their parents’ genomes, both history and genetic linkage create patterns in gene frequencies between populations. These patterns are related to health outcomes, like COVID-19, via structural injustices in society and thus can potentially lead to confounding bias in studies of blood group and COVID-19. In addition to these confounding pathways, our DAG also identifies several potential sources of bias which could arise if non-confounders are adjusted for or restricted on in the analysis, particularly pre-existing ABO blood type and blood bank as a source of controls.

### Common issues in ABO blood group / COVID-19 studies

Overall, we identified several common methodological issues in the data collection or in the analysis procedure which could decrease the internal validity and/or the generalizability of results and lead to inaccurate or misleading conclusions. We briefly summarize the most common threats to validity, including confounding, selection bias, and collider bias.

#### 1. Uncontrolled confounding

Confounding is one of the most common threats to validity in observational research. Although ABO blood type is genetic, there are a number of potential confounding pathways that need to be considered. For example, in our DAG the pathway ABO blood group <-historical injustice -> structural racism-> occupational exposure -> COVID represents a potential mechanism by which ABO blood group could be found to be associated with COVID even when blood group has no *biological* relationship to COVID risk or severity. Studies of the relationship between ABO blood group and COVID which do not adequately control for the impact of structural racism may find a positive association between blood groups which minoritized groups have higher frequencies and COVID incidence and/or severity. This is bias may affect the findings in 9 (37.5%) out of the 24 studies, including Abdollahi et al., Barnkob et al., Boudin et al., Ellinghaus et al., Hoiland et al., Niles et al., Taha et al., Zalba et al., and Zietz & Tatonetti. When data on history, structural racism, or their proxies (such as race/ethnicity or socioeconomic status) are unavailable, alternative adjustment sets should consider including COVID-19 exposure risks, such as occupational exposures, as well as factors that could affect access to testing, medical care, and/or hospitalization.

In addition to uncontrolled confounding pathways, a number of studies inappropriately adjust for variables which are not likely to be confounders. 6 (25%) out of the 24 identified articles adjusted for mediators in their analysis, including variables such as chronic medical conditions, medications, as well as physical and behavioral characteristics, could be consequences of blood type if ABO blood group has any biological impact on health. Since these variables may also impact COVID risk or severity, controlling for these potential mediators does not address confounding, and could even introduce bias to an analysis (Aschengrau & Seage, 2020, p. 210).

#### 2. Bias in sample selection

14 (58.33%) of the studies we identified relied on prior ABO blood typing to determine study eligibility (denoted by the box around prior ABO typing on the DAG in Figure 2). However, having an existing record of ABO blood type could itself be associated with ABO blood type in two ways. First, historic and structural injustices may mean individuals in certain areas or populations have less access to care and therefore be less likely to have their blood type recorded prior to contracting COVID-19. In addition, if ABO blood type is associated with any (non-COVID) health conditions, either directly or indirectly via linkage disequilibrium with other genetic loci, individuals with particular ABO blood types may be more likely to have chronic health conditions, and thus have more frequent interactions with the healthcare system resulting in more opportunity for a prior blood type record to exist.

Since health status and access to care both affect the likelihood that an individual’s blood type is available in their medical record, we call prior ABO typing a common effect, or collider. When a variable is a collider, restricting on it will typically create bias (Griffith et al. 2020). Thus, we expect any studies which select their study sample based on prior ABO blood type availability could detect a statistical association between at least one blood type and COVID, *even if blood type is completely unrelated to COVID risk or severity*. This bias could occur if there are direct causal relationships between ABO blood types and any health conditions (such as have been proposed for cardiovascular diseases, cancer, and metabolic diseases (Abegaz, 2021; Groot et al., 2020; Mandato et al., 2017)), or if there are any common causes of ABO blood group and health status (i.e. confounding variables) such as might result from linkage disequilibrium between ABO blood type and another important health-related gene (e.g., U1).

#### 3. Bias in control selection

Of the 12 case-control studies we identified, 4 (25%) selected control information by obtaining the population distribution of ABO blood type from blood bank records before 2020. This is an attractive control selection strategy, since it does not require identifying specific control individuals. However, it is likely that this strategy introduced bias. In case-control studies, the fundamental tenet of control selection is that controls must be selected independently of their exposure status (here, blood type). Unfortunately, it is well-known that blood banks intentionally over-recruit individuals with type O blood due to their “universal donor” status (Dzik et al., 2020; The American National Red Cross, n.d.). Therefore, it is expected that blood banks would have *higher* frequencies of type O blood than the general population. We should thus also expect that studies using a blood bank-based control group would always find that the frequency of type O blood is lower among COVID cases than in the blood bank control sample. Such an association would be spurious and not rely on any real, biological, relationship between blood type and COVID.

Of the remaining case-control studies, 6 (50%) selected controls from hospitalized patients with other diseases, and 2 (12.67%) selected controls in the same hospital with COVID cases around the same time (Dzik et al., 2020; Fan et al., 2020). This is generally a reliable control selection process, but it is not guaranteed to be free from bias. Controls should be selected to represent individuals who, if they developed the outcome, would be included in among the cases. For many diseases, patients in the same hospital adequately represent people who would be recruited as cases and this may also be true for COVID-19 but studies typically did not justify their choice of control group.

#### 4. Failure to address missing data

Another issue prevalent among the studies is the absence of discussion of missing data. 16 (66.67%) of the 24 studies did not address how they dealt with missing data. It is unknown whether the exclusion of participants are similar across ABO blood types (Sterne et al. 2016), and we cannot control for the effect of missing data without more information on handling of missing data. 14 (58.33%) of the articles restricted participants based on previous blood type information, with an average of 28% of subjects excluded due to missing blood type data. Excluding individuals without prior ABO blood type information could exacerbate selection bias if individuals without this information are systematically more likely to have lower access to medical care, or be members of minoritized or underrepresented groups (Aschengrau & Seage, 2020, p. 210).

#### 5. Inappropriate statistical analysis

ABO blood group is a four-level nominal categorical exposure variable. When attempting to identify whether any particular level of a categorical variable is at higher or lower risk of an outcome, it is important to use a two-step hypothesis testing procedure. This process would first conduct an overall test of homogeneity, where the null hypothesis is that there is no difference in the outcome probability among any of the levels of the exposure. If, and only if, this null hypothesis is rejected, it is appropriate to then conduct multiple two-way comparison tests to assess which exposure levels have higher or lower outcome probabilities. Failure to follow this procedure can dramatically increase the likelihood of falsely concluding that exposure categories differ in outcome frequency. Despite this, 16 (66.67%) articles failed to report an overall result of the chi-square test among the blood groups and it is unclear whether they conducted any overall first stage testing.

#### 6. Insufficient sample size

Small sample size is another important factor to consider when evaluating study quality and can increase the risk of chance findings or decrease the ability to control for confounding or other biases. Since ABO blood group has 4 categories, the required sample size is larger than would be required for a binary exposure. 2 of the 7 (29%) cohort studies used a total sample size less than 400, as did 4 of 12 (33%) case control studies. Small sample size could make assessing COVID-19 outcomes especially challenging for people with Type AB blood groups, since this blood group is relatively rare (Garratty & Glynn, 2004). The average proportion of Type AB subjects was 6.5% for cases and 5.8% for controls in case control studies, while the average proportion of Type AB subjects was 3.8% for cohort studies. Small sample size could amplify selection bias when there are losses to follow-up or when there is self-selection of participants into the study (Aschengrau & Seage, 2020, p. 293). Though there is no definite criteria for the minimum sample size, it would have been helpful for authors to discuss how the sample size was determined based on a meaningful effect size and desired statistical power.

#### 7. Potential for misclassification

Finally, most studies did not include sufficient data for us to assess the potential for bias due to misclassification. However, it is reasonable to expect some measurement error in the outcomes for at least some studies, because COVID-19 diagnostic tests may return false negative results (Dinnes et al., 2020). Measurement error in the outcome may impact the precision of results (Aschengrau & Seage, 2020, p. 317). However, it seems reasonable to expect measurement error in outcomes are independent of ABO blood type and thus any outcome measurement error would likely be non-differential with respect to the exposure.

Measurement error in the exposure is generally unlikely for studies using individual-level patient data but could be a problem in the ecological studies. These studies both used non-academic, non-governmental websites to collect data on ABO blood group distributions for the countries they included. While it seems unlikely that error in ABO blood group distributions would be related to COVID-19 outcomes, the exposure is a categorical variable and so even non-differential measurement error could create spurious associations. Finally, 4 (16.67%) of the articles performed their analysis by dichotomizing the four ABO blood groups, for example by comparing Type O blood groups to Non-O groups. This analytic approach effectively builds-in measurement error, since there does not appear to be a biological justification to combine blood types (Aschengrau & Seage, 2020, p. 286).

## Discussion

Overall, our review of the evidence suggests there is no true relationship between ABO blood type and COVID-19 infection, severity, or mortality. Certainly, there is no evidence to support the conclusion of the *existence* of a biological relationship. We recommend that future efforts to identify groups at high- or low-risk of COVID morbidity and mortality refocus on other potential causal factors. However, since many other potential causal factors will encounter similar challenges in identifying and estimating causal effects, we also urge caution when designing studies to assess COVID-19 morbidity and mortality risk, especially those related to biomarkers which cannot be experimentally modified (Tennant & Murray 2020).

We have described 7 potential sources of bias that may have created spurious associations between ABO blood type and COVID risk or severity – including confounding, restriction on a collider (a bias commonly known as ‘selection bias’ or ‘collider bias’), inappropriate control selection, missing data, inappropriate statistical analyses, insufficient sample size, and misclassification. Confounding is a common problem in observational studies, but many researchers may wrongly conclude that the genetic nature of ABO blood type means it cannot be confounded due to confusion about the difference between individual- and population-level causes of genetics. Selection, or collider, bias is somewhat less well-known outside of the epidemiology and economics communities, but the role of this bias in COVID studies has been extensively discussed (Griffith et al., 2020).

Of note, all the potential biases we identified could be remedied at the study design phase. Control individuals should be enrolled independently of their blood type and all study participants should have their ABO blood type measured for the current study. These two precautions would prevent three of the potential confounding or selection bias pathways from causing bias in the study results (Ellinghaus et al., 2020; Sardu et al., 2020). The remaining pathway – confounding via the impact of historic injustice and structural racism – could be controlled by collecting and adjusting for all aspects of these variables which increase COVID risk. This is not easy to do, but at least one of the studies we identified was likely successful in reducing the impact of this confounding due to their choice of sample population (Boudin et al., 2020). This study focused on French airmen who shared a common exposure aboard an aircraft carrier. Due to shared occupation and employer, these individuals will have had similar access to care and similar occupational exposures, as well as being within a narrow range of age, gender, and physical health / fitness, greatly reducing the potential for bias due to the impacts of historic factors or structural racism on COVID-19 risk and outcomes.

As we finalized our manuscript, a large case-control study was published in JAMA Open (Anderson et al, 2021). This study was well-done but did repeat the error of restriction based on prior blood type data availability. Although there was limited control for confounding, the study was conducted in a homogenous sample of patients in Utah, Nevada, and Idaho so the potential for bias is likely limited. Anderson et al found no association between ABO blood type and any COVID outcomes.

Finally, it is worth noting that the hypothesis that SARS-CoV-2 or COVID-19 may be related to ABO blood type appears to derive from a single research letter in JAMA detailing an analysis of 45 staff at the Prince of Wales Hospital in Hong Kong during the 2003 SARS outbreak. This study did not conduct a test of the global null hypothesis but reported that staff with Type O blood group were less likely to have contracted SARS. The research letter includes the raw data table and based on that we estimate that the chi-squared for the global null hypothesis is 3.7 giving a p-value of 0.3 and failing to reject the null hypothesis that the distribution of ABO blood type was identical between staff members who contracted SARS and those who did not.

In conclusion, we believe that the evidence does not support the claim that COVID-19 risk is determined by ABO blood group. We urge clinicians, public health officials, and science communicators to council patients and the general public to take infection control precautions regardless of their ABO blood group.

## Data Availability

Data extraction tables are available upon request.

## Acknowledgements

This work was supported by the Epidemiologic COVID-19 Response Corps at Boston University School of Public Health.

## Appendix 1: Risk of bias assessment for each article

**<u>Abdollahi et al.’s</u>** study assessed the association between ABO blood group and susceptibility to COVID-19 among patients at the Imam Hospital Complex in Iran. They reported an increased risk in patients with Type AB and a decreased risk in patients with Type O blood. Despite using stratifications by sex, the study has not adjusted for the influence of historic or structural racism factors, nor proxies such as race or ethnicity. The Iranian population is not ethnically or culturally homogenous, and thus the potential for confounding related to historic or ongoing oppression exists. Thus we conclude that this study has a moderate bias due to confounding according to the ROBINS-I tool. Cases and controls were both selected from hospitalized patients at the one chosen medical center, but it is unclear from the article whether controls were identified from blood bank participants prior to 2020 or from outpatient and inpatient non-COVID patients during 2020. As a result, we list the potential for bias due to selection of participants into the study as moderate. Finally, the authors did not discuss how they handled missing data, leaving the possibility of bias due to missing data unclear. Classification of exposure, measurement of outcomes, and choice of outcome were all judged to be low risk of bias. *Overall, the study has a moderate level of bias due to potential confounding and selection bias*.

**<u>Ahmed et al.’s</u>** study assessed COVID-19 susceptibility among women enrolled at maternal units in the UK. They report an increased risk of asymptomatic infection among pregnant women with Type A blood group, and a decreased risk among pregnant women with Type O blood group. However, this association appears to be entirely driven by findings in the 208 Black, Asian, and Minority Ethnic (BAME) women, with no difference in infection risk by ABO blood group among the 146 white European women. The stratified analysis suggests that confounding bias may be present in the study results. The study sample included all pregnant women presenting to two participating hospitals, and ABO blood type was collected at the time of COVID-testing. This reduces the likelihood of bias in selection of participants but introduces the potential for correlated measurement error in exposure and outcome variables. The authors do not address potential measurement error, nor do they address the issue of missing data. Finally, given that this is a cross-sectional study that did not recruit based on outcome status, it is unclear why the authors chose to assess ABO blood group distribution by COVID status rather than assessing the frequency of COVID infection within each blood group. A crude assessment based on their reported data indicates that the choice of analysis affected the study conclusion. *Overall, the study has a moderate overall risk of bias*.

**<u>Alkout and Alkout’s</u>** ecological study assessed the relationship between ABO blood type distribution and risk of detected SARS-CoV-2 infection globally. Results of such an ecological study might not accurately show individual level association between blood types and risk of infection (Piantadosi, Byar, & Green, 1988). The authors did not adjust for any confounders, leading to serious risk of bias due to confounding. Moreover, there might be moderate bias in selection of participants and in classification of interventions because they only included countries with COVID data reported on the Worldmeter website and ABO blood group distributions reported on Wikipedia. There is also a high possibility of bias in the measurement of outcomes due to inconsistencies in reporting and testing criteria between countries. The only adjusted analysis the authors report is on data stratified by continent. However, there is no clear rationale for this adjustment. *Overall the study has serious risk of bias*.

**<u>Anderson et al.’s</u>** case control study among 107,796 participants investigated the association between ABO blood types and COVID infection risk as well as severity after infection. This study was published after our systematic review phase. The authors found no association between ABO blood types and COVID outcomes in their relatively large sample selected from electronic health records in Utah, Idaho and Nevada. Age, sex and Rh factors were adjusted for in the analysis of potential association. The authors discussed the association between non-white race and testing positive, and report no change in results when restricting the study population to white individuals only. The study recruited participants based on health records of COVID test results, but limited their sample to those having an existing blood type record only. The authors do not report the percentage of participants excluded due to missing blood type record, and the study thus has a moderate risk of bias due to selection of participants. The authors do not address the issue of missing data and have an unclear risk of bias in this domain. A test of the global null hypothesis was reported before reporting the results of pairwise comparisons between blood groups. *Overall this study has a low to moderate risk of bias*.

**<u>Ansari-Lari & Saadat’s</u>** ecological study was conducted in 86 countries to assess the relationship between ABO blood group or Rh-type and COVID outcomes, including detected cases and mortality. The authors consider potential confounding by life expectancy, gross national income, medical care, and tobacco smoking, but use an automated model building procedure (backward selection) to determine the final set of covariates included and do not specify which if any were excluded from the final model. The risk of bias due to confounding is thus moderate. The study also has a moderate bias in selection of participants. Similar to Alkout & Alkout, this study only included countries with COVID cases listed in Worldometer, and obtained exposure data from Rhesusnegative.net but do not discuss how countries were selected for inclusion in this list. Additionally, potential bias in classification of interventions might exist, as the blood type distribution information was collected from an unofficial website which lacks citations. This study might also have a serious bias in the measurement of outcomes because it does not discuss the impact of different testing and reporting criteria between countries. Furthermore, the authors compared only Type A and Type B blood groups with no justification for this choice or overall test of the global null hypothesis. There is therefore potential for serious bias in the selection of reported results. Finally, the authors did not discuss handling of missing data. *Overall the study has serious risk of bias*.

**<u>Arac et al.’s</u>** article is a case-control study on the association between ABO blood types and COVID hospitalization conducted in Turkey. The authors reported finding no association. The authors did not adjust for any confounders, nor do they provide a rationale for not doing so; we conclude that this study thus has a serious risk of bias due to confounding. Similar to Abdollahi et al., the study only selects cases from patients admitted to one hospital. Although suspected patients are also admitted to the hospital, the article did not specify how these suspected patients were identified and it is likely that asymptomatic or mild infections are excluded from the study. There is potential for moderate risk of bias in selection of participants if there is any relationship between ABO blood group and access to testing or care. The authors do not report an overall test of the global null hypothesis that the blood types are distributed independently of COVID status. Finally, the authors do not address the issue of missing data and have an unclear risk of bias in this domain. *The study has a serious overall risk of bias*.

**<u>Barnkob et al.’s</u>** study is a retrospective cohort analysis of all Danish individuals who were tested for SARS-CoV-2 with previously recorded ABO and RhD blood groups. The main analysis does not adjust for any confounders, but the authors report a sensitivity analysis adjusting for immigrant status among individuals of non-Western origin. The risk of bias due to confounding appears to be low due to the negligible change between main and sensitivity analysis results. However, the study has a serious risk of bias due to selection of participants into the study, since only 473,654 individuals (56%) had known blood group types among 841,317 people who were tested for SARS-CoV-2 during the study period. *Overall, the study has a serious risk of bias primarily attributable to the potential for selection bias*.

**<u>Boudin et al.’s</u>** study is a retrospective cohort study conducted among crewmen of a French navy aircraft carrier. The authors report no association between ABO blood group and COVID infection. The authors did not adjust for confounding, but due to the nature of the study sample we judge the risk of bias due to confounding to be low. In addition, while the study population is unique in important ways (young, healthy, largely male), very few individuals were excluded from the study sample so the risk of bias due to selection of the study population is low. Note, that the choice of study sample may restrict the applicability of study results to other populations. *Overall, this study has a low risk of bias but relevance of results to other settings should be assessed*.

**<u>Dzik et al.’s</u>** article reports on a case-control study of COVID-19 patients at Massachusetts General Hospital and Brigham and Women’s Hospital. The authors compared ABO blood groups between patients who did and did not survive following hospital admission for COVID-19 from Feb 12 to May 13 2020. They found no significant association between ABO blood group and COVID-19 mortality in an overall test of the global null hypothesis. The authors do not report any adjusted analyses, and there is therefore potential for bias due to confounding. In particular, the authors report that the hospitalized COVID-19 patients are somewhat more likely (although not statistically significantly) to have Type O blood than other hospitalized patients at these two hospitals. However, since all study participants were hospitalized for COVID-19, the potential confounding pathways are reduced (for example access to care is unlikely to be a confounder here). There is a moderate risk of bias in selection of subjects since the authors restrict their sample to individuals with an existing ABO blood type record. 65 % of hospitalized patients had ABO testing performed, suggesting 35% were excluded for missing blood type data. *Overall, we determined that this study has a moderate risk of bias*.

**<u>Ellinghaus et al.’</u>**s study reported on a genome-wide association study (GWAS) conducted at seven hospitals in the Italian and Spanish epicenters of COVID. The authors adjusted for age and sex but do not consider other confounders. They reported an increased risk in participants with Type A and a decreased risk in participants with Type O blood. Based on our assessment of the literature, it seems unlikely that age or sex would confound the association between ABO blood group and COVID outcomes. They did adjust for multiple hypothesis testing in the GWAS by using a p< 5×10^−8^ threshold. The study sample consists of patients hospitalized with severe COVID, and control participants recruited from blood donor clinics plus pre-existing panels of genomic data. The study has a moderate potential for bias due to selection of participants because it is unclear whether the individuals included in the control group would have been admitted to one of the seven study hospitals if they developed severe COVID. The authors did exclude 25 controls due to missing covariate (age, sex) data. *Overall, this study has a moderate risk of bias*.

**<u>Fan et al.’s</u>** case-control study was conducted at Zhongnan Hospital of Wuhan University in China from January 1, 2020 to March 5, 2020. The study reported that patients with Type A, especially females, are more likely to be infected. The study included a total of 105 COVID-19 cases and 103 controls, which could be statistical concerns of small sample size. The study only adjusted for gender and thus has a moderate risk of bias due to confounding. The study also has a serious risk of bias in selection of participants, since the control participants were subject to a wide range of exclusion criteria not applied to the case participants, including no history of respiratory infection, no other infectious diseases, and no severe liver or kidney dysfunction. The study did not mention any information of missing data. *Overall, the study has a serious risk of bias*.

**<u>Hoiland et al</u>**. conducted their study in 6 metropolitan hospitals in Vancouver, Canada. They compared the distribution of ABO blood groups (dichotomized as anti-A antibody vs no anti-A antibody) in patients admitted to the intensive care unit (ICU) for COVID-19 with the distribution of ABO blood group in historic blood bank data. The sample size of the study is relatively small, with only 95 patients in total after excluding those with missing ABO blood type. The study has a moderate risk of bias due to confounding – the authors assess a range of clinical presentations and outcomes but do not adjust for any covariates in their comparison of ABO blood group distribution. The study also has a serious risk of bias in selecting participants because around 20% of patients were excluded due to lack of ABO blood group data. *Overall, the study has a serious risk of bias*.

**<u>Latz et al</u>**. report on a multi-institutional study of all adults with a positive COVID-19 test result recorded at any of five hospitals in Massachusetts, USA, from March 6 to April 16 2020. The analysis adjusted for a range of confounding factors such as sex, primary language, race, and Rh phenotype. However, the authors also adjusted for a range of health conditions which could be either confounders or mediators, including aspirin use, calcium channel blocker use, and history of chronic kidney disease, coronary artery disease, stroke or diabetes mellitus. The authors do not discuss why they believe these are not mediators, and used an automated variable selection procedure for choosing adjustment variables which is not guaranteed to select confounders. Based on this, we conclude there is a serious potential for bias due to confounding. There is serious potential for bias in selection of participants due to the reliance on existing ABO blood group records which were only available for 1289 (17%) of 7684 individuals with positive COVID-19test results. *Overall this study has a serious risk of bias*.

**<u>Leaf et al.’s</u>** study used the data from the Study of Treatment and Outcomes in critically ill patients with COVID-19 (STOP-COVID) cohort collected at 67 hospitals across the United States. The study sample includes adults admitted to participating ICUs with laboratory-confirmed COVID-19. The authors adjusted for race/ethnicity, which could be sufficient to control for confounding if adequately measured. However, there is serious risk of bias due to selection of study participants since approximately one-third of potential patients were excluded due to missing ABO phenotype data. In addition, the authors compared the distribution of ABO blood type in their cohort with blood bank donors in the US. Despite using a cohort to select cases, this study is therefore a case-control study. *Overall, the study has a serious risk of bias*.

**<u>Li et al.’s</u>** retrospective case-control study is based on patient data in Wuhan, China. This study adjusted for sex but not race or ethnicity. However, given the study location, this might be sufficient for limiting confounding. Cases were individuals diagnosed with COVID-19 who died or were discharged between Feb 1 and March 25 2020, and controls were selected from existing data and published papers. The potential for bias in selection of participants is moderate since the control data comes from different hospitals than the case data. The authors do not report an overall test of the global null hypothesis, and this may affect their results. *Overall, the potential for bias is moderate*.

**<u>Nile et al.’s</u>** study reports on a nationwide cohort study of detected COVID-19 infection among women using electronic medical record data from the USA. Analyses were stratified by age, and by race/ethnicity. However, nearly 70% of the patients were excluded from the analysis which adjusted for race/ethnicity due to clinician-reported, rather than self-reported, data. The study compared women with positive versus negative SARS-CoV-2 test results. There is moderate potential for bias due to selection of study participants. Although the indication for testing was similar across the US during the study period (May-June 2020), participants with negative test results may be more likely to work in occupations where regular screening tests were required or provided. *Overall, the risk of bias is moderate*.

**<u>Padni et al.’s</u>** ecological study assessed patterns of COVID-19 mortality status across 28 states and 8 union territories in India and association with ABO distributions in these states. There is serious potential for bias in classification of interventions since data on ABO distributions were obtained from a literature search and no assessment of study quality was provided. In addition, states for which no previous reports of ABO blood type distribution were identified were excluded from the analysis. There is also a potential for bias due to confounding – the authors do not report any adjusted analyses. *The article has a serious overall risk of bias*.

**<u>Sardu et al.’s</u>** prospective cohort study investigates the association between ABO blood group and prognosis among hypertensive COVID-19 patients in an Italian hospital. The sample size for this study is relatively small (162 participants) and the authors had a large list of exclusion criteria which may limit the generalizability of their study. ABO blood type was dichotomized as Type O or non-Type O with no test of the global null hypothesis. The authors consider a large number of covariates but many of these are health status variables that could potentially be mediators, and they choose the final set of analytic variables based on an automated selection procedure. The risk of bias due to confounding is therefore moderate. *Overall this study has a moderate risk of bias and the applicability beyond the very specific patient population is unclear*.

**<u>Taha et al.’s</u>** case-control study was conducted in Sudan using an online survey sent to individuals with confirmed COVID-19. Survey participants self-reported blood type, COVID-19 symptoms, prior history with malaria and chronic disease. Control data were extracted from maternity hospital medical records of 1000 healthy volunteers. The potential for bias in sample participant selection is high, since it is unclear how survey participants were identified. The risk of bias in misclassification of intervention is also serious, because the study used self-reported blood groups for cases but hospital records for controls. Thus, measurement error in ABO blood type likely differs between cases and controls. Finally, the authors do not report any adjusted analyses and the potential for confounding bias is unclear. *Overall, this study has a serious risk of bias*.

**<u>Wu et al.’s</u>** retrospective case-control study analyzed 187 patients of the Third Xiangya Hospital of Central South University and the First Hospital of Changsha, in Hunan province, China, between January 20, 2020 and March 5, 2020 to explore the blood group distribution and clinical characteristics among those patients. The authors discuss age, sex, and comorbidities but do not adjust for any potential confounders other than restricting to Han Chinese ethnicity. However, this restriction may be sufficient to control for confounding in this setting. Control data were obtained from hospital Han Chinese patients with existing ABO phenotype and no known COVID-19 infection between Jan 2019 and Feb 2020; cases were all Han Chinese patients hospitalized or discharged from the study hospitals between Jan 20, 2020 and March 5 2020, with known ABO phenotype. The authors do not report the number of individuals excluded for missing ABO blood group information. There is also potential for the control group to include individuals with undiagnosed COVID since the control period and case period overlap, and there was no restriction on the types of hospital admissions considered for control participants. Finally, the authors do not report a test of the global null hypothesis. *Overall, this study has serious potential for bias, largely due to lack of clarity in sample selection*.

**<u>Yaylaci et al.’s</u>** article is a cross-sectional study on the prognosis of COVID-19 among 397 COVID-19 patients in Turkey between March 19 and April 19 2020. The authors compared ICU admission and mortality by ABO blood group among individuals with COVID-19 but provide minimal details on the study sample. They do not report whether all individuals were phenotyped for ABO blood group or whether there were any participant exclusions. In addition, they do not report any control for confounding nor do they report an overall test of the global null hypothesis. *Overall, we judge this study to have a serious risk of bias due to a lack of available information to better assess the study quality*.

**<u>Zalba Marcos et al.’s</u>** article compares the distribution of ABO blood types between patients hospitalized at 2 public hospitals in Navarra, Spain with a donor and transfusion database. The study has a low risk of bias due to confounding. The authors adjusted for age and sex, and the demographics of Navarra suggest that race/ethnicity adjustment may not be warranted. There is low to moderate risk of bias in sample selection due to the use of database controls, but this appears to include transfusion recipients not just donors which may be more representative of the general population. The authors report a global null hypothesis test. *Overall, we judge this study to have a moderate potential for bias based on the reported information*.

**<u>Zhang et al</u>**. report a case-control study on COVID-19 patients in the ICU in Wuhan Jinyintan hospital from December 2019 to February 2020. They compared ABO blood types between survivors and non-survivors during the study period. The authors do not report any confounding adjustment, but it is unclear whether this is a potential source of bias given that these data are from very early in the pandemic. The sample size is small (134 patients) and the authors assess several outcomes. There is thus potential bias in the choice of reported results. The authors also do not report a test of the global null hypothesis. An additional analysis compares ABO blood group in COVID-19 cases with the general population of Han Chinese from existing data sources; this analysis may be subject to moderate bias due to study participant selection. *Overall, this study has a low risk of bias for the primary outcome of mortality, and moderate risk of bias for all other outcomes and comparisons*.

**<u>Zhao et al.’s</u>** case-control study was conducted in China and reports on data from Wuhan and Guangdong. The authors discuss the potential for confounding by age and gender but cannot adjust for gender due to missing data. It is unclear how the cases were selected. However, the authors do report collecting case ABO phenotype for the current study. Control ABO phenotype distribution relied on published estimates which might not be appropriate. The authors do not conduct a test of the global null hypothesis. *Overall, the risk of bias in this study is serious primarily due to a lack of information on study samples and confounding*.

**<u>Zietz & Tatonetti’s</u>** case-control study was conducted at the New York Presbyterian hospital in the US. However, they do not discuss the criteria for case selection. Control data were obtained from historical medical records, but excluding anyone who tested positive for COVID-19 in the study period. This introduces potential for bias in the sample selection process. The study adjusted for age, sex and chronic conditions using multivariate logistic regressions were used. The authors do not report adjustment for race/ethnicity, and do not discuss whether any of the chronic conditions could be mediators. Eligible patients with no blood type record or multiple contradictory blood type results are excluded. The authors did not discuss missing data. *The article has a serious overall risk of bias*.

